# Real-world comparative outcomes of GLP-1 RA and semaglutide prescription among individuals with type 2 diabetes

**DOI:** 10.1101/2025.06.03.25328908

**Authors:** Maxwell Salvatore, Bingyu Zhang, Huilin Tang, Yiwen Lu, Dazheng Zhang, Ting Zhou, Yuan Lu, Anastassia Amaro, Marylyn Ritchie, Yong Chen

## Abstract

Glucagon-like peptide-1 receptor agonists (GLP-1 RAs) are increasingly prescribed for type 2 diabetes (T2D) and weight management, yet their broad health impacts in real-world settings remain understudied. Using data from the All of Us Research Program (n=18,746), we conducted both intention-to-treat and per-protocol phenome-wide association studies comparing diagnoses following GLP-1 RA prescription, including semaglutide-specific analyses, to those following sodium-glucose cotransporter-2 inhibitor (SGLT2i) and dipeptidyl peptidase-4 inhibitors (DPP4i) prescriptions in individuals with T2D. GLP-1 RAs were associated with reduced risks of genitourinary and dental conditions relative to comparators, while semaglutide was linked to lower risks of cardiac arrhythmia, hyperglycemia, and chronic kidney disease. However, GLP- 1 RAs were also associated with increased risks of dysthymic disorder and vitamin D deficiency. Time-to-event analyses revealed modest delays in diagnosis for key outcomes. These findings underscore differences in downstream associations across second-line T2D therapies and highlight semaglutide’s distinct profile. Results may inform clinical decision-making and motivate further research on effectiveness, safety, and personalized prescribing.

## 1. MAIN

Type 2 diabetes mellitus (T2D) is a pervasive chronic condition that affects millions globally, with devastating complications when poorly managed.^1–3^ Recent years have witnessed the emergence of innovative therapies that have been crucial in improving glycemic control and patient outcomes.^4,5^ Among these therapies, glucagon-like peptide-1 receptor agonists (GLP-1 RAs) have revolutionized the diabetes management landscape.^6–9^

Initially developed for T2D management, GLP-1 RAs have garnered substantial attention since the introduction of exenatide (Byetta, Bydureon) in 2005. These drugs mimic the action of the incretin hormone GLP-1, inhibiting glucagon release and slowing gastric emptying, thereby aiding in glucose regulation. The FDA approved semaglutide (Wegovy) for chronic weight management in 2021, marking a significant expansion of GLP-1 RA use beyond glucose control.^10^ Semaglutide is noteworthy for research due to its superior efficacy (15% weight loss within one year versus 5-10% with earlier ^11^–^16^), unique pharmacokinetic profile resulting from its modified molecular structure,^17^ and rapid adoption, particularly among populations without diabetes.^13^ This approval has sparked enormous public interest and scrutiny in GLP-1 RA use and its clinical effects beyond glycemic control. Concurrently, telehealth platforms dramatically expanded access to these medications,^18–21^ with some services explicitly marketing semaglutide for weight loss in non-diabetic populations. This rapid adoption through novel care delivery models underscores the urgent need for robust safety surveillance and comparative effectiveness research, including ones specifically focused on semaglutide.

Despite their growing popularity^22–25^ (e.g., GLP-1 RA prescriptions increased, 3% between September and December 2023, 21% between December 2023 and March 2024, 8% between March and June 2024, and 12% between June and September 2024^26^) and the media’s portrayal of expected and unexpected benefits of GLP-1 RAs,^27,28^ there exists a paucity of real-world data examining their downstream health effects. Current literature primarily focuses on clinical trial outcomes,^12,13,16,29–31^ leaving a substantial gap in our understanding of the real-world impacts of these medications. Understanding the potential benefits and, more importantly, any associated harms of GLP-1 RA use is crucial.

Because GLP-1 receptors are present in the pancreas, lung, heart, kidney, intestine, skin, and several regions of the central nervous system,^32^ GLP-1 RA studies have considered widespread impacts, including impacts on cardiovascular disease,^33–38^ kidney disease,^33,35,37–39^ neurocognitive diseases,^40–43^ and substance use disorders.^44–48^ Using the US Department of Veterans Affairs databases, Xie, Choi & Al-Aly (2025)^49^ systematically tested for associations across 175 clinical outcomes comparing GLP-1 RA prescription to three anti-diabetes therapies among individuals with T2D. It corroborated many studies, finding GLP-1 RA use associated with reduced risk for substance use^44–48^ and psychotic disorders, seizures,^50,51^ neurocognitive disorders,^52–54^ coagulation disorders, cardiometabolic disorders,^33–38^ infectious illnesses, and several respiratory conditions. Importantly, their study also found *increased* risk of gastrointestinal disorders,^55–57^ hypotension and syncope (some prior evidence of blood pressure-lowering^58–60)^, arthritic disorders, nephrolithiasis (alternative medications may be relatively more protective^61,62^), interstitial nephritis, and drug-induced pancreatitis (adverse events have been reported^63,64^) associated with GLP-1 RA use. Systematic, phenome-wide association studies (PheWAS) are hypothesis-generating and are crucial for identifying knowledge gaps and generating information.

Our study extends beyond the work of Xie and colleagues by examining an even broader range of clinical outcomes (up to 1,026 phenotypes) while leveraging electronic health record (EHR) data from the All of Us Research program (AOU),^65^ a nationally recruited EHR-linked biobank specifically designed to oversample groups historically underrepresented in biomedical research. This diverse cohort provides a significant opportunity to validate previous findings in a demographically different population from the predominantly older, White, male veterans studied by Xie and colleagues.

By examining real-world usage patterns and health outcomes, we seek to explore the downstream phenotypes associated with GLP-1 RA use. Specifically, this study compares the phenotypic consequences of GLP-1 RA prescription with those of other second-line anti-diabetes therapies, namely sodium-glucose cotransporter-2 inhibitors (SGLT2i) and dipeptidyl peptidase-4 inhibitors (DPP4i). To achieve these objectives, we employed a robust methodological approach combining phenome-wide propensity-score matched intention-to-treat (ITT) and per-protocol (PP) Cox proportional hazard models, alongside restricted mean survival time (RMST) analyses. The ITT analysis preserves randomization by analyzing participants according to their initial treatment assignment regardless of adherence. In contrast, the PP analysis considers only those who adhered to the treatment, providing complementary perspectives on prescription impacts. This approach allows for evaluating relative and absolute associations between GLP-1 RA prescription and a wide array of health outcomes in a demographically diverse population.

## 2. RESULTS

### 2.1. Descriptive evaluation of the data

The analytic cohort comprised 18,746 adults with T2D who received a qualifying drug prescription on or after January 1, 2018 (Figure 1). They had an average age at T2D diagnosis of 53.0 (standard deviation 11.8) years, were 57% female, were 49% non-Hispanic White, 62% were with obesity (BMI ≥ 30), had an average of 13.3 (8.2) years of EHR follow-up, and an average Quan-weighted Charlson Comorbidity Index (CCI) score of 2.8 (2.6) (Table 1). These individuals were grouped into three cohorts based on drug classes: GLP-1 RA (n=8,798), SGLT2i (n=5,111), and DPP4i (n=4,337).

**Figure 1.**
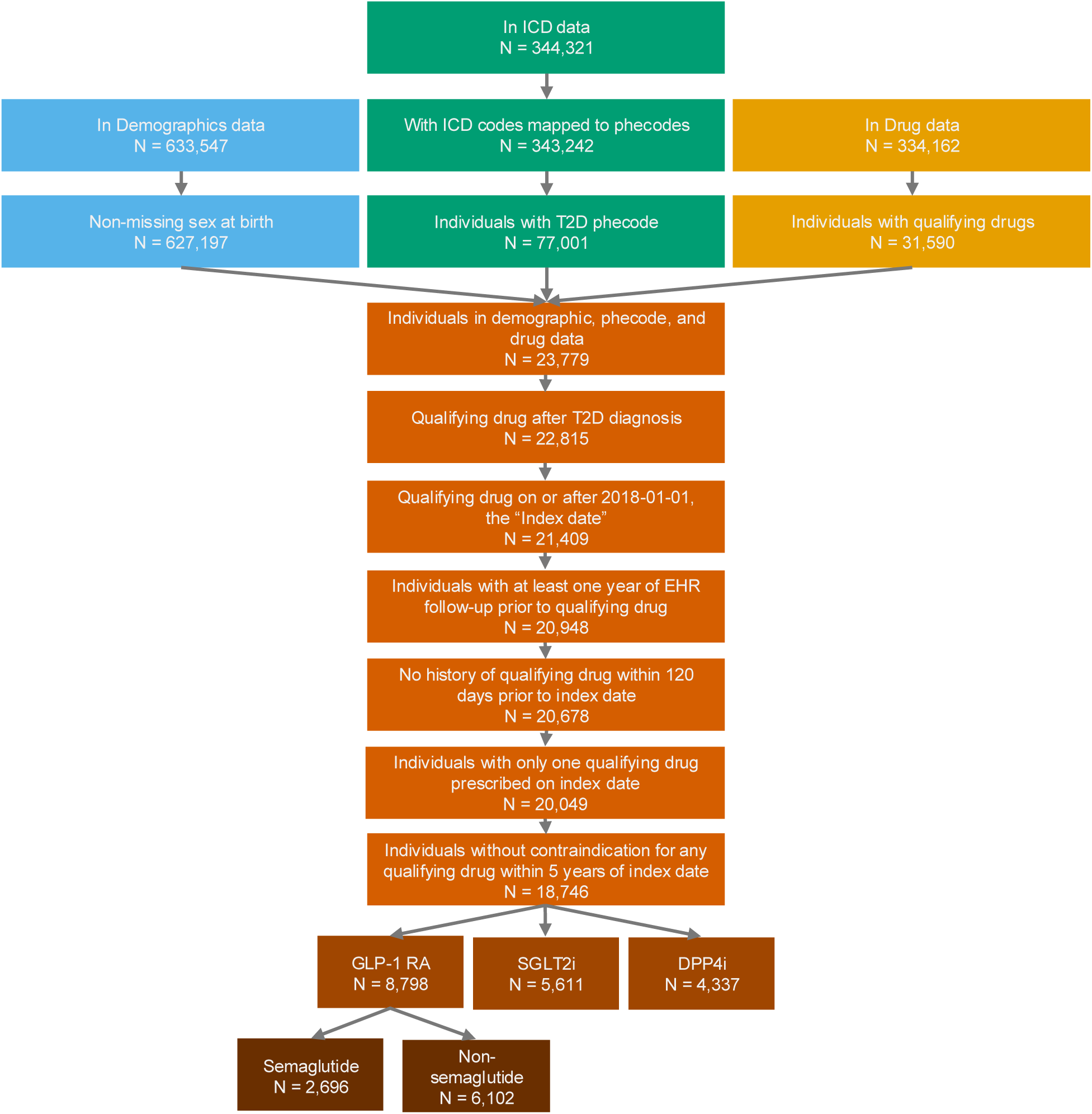
Flowchart depicting sample sizes throughout thresholding process. Abbreviations: dipeptidyl polypeptidase-4 inhibitor; GLP-1 RA, glucagon-like peptide-1 receptor agonist; sodium glucose cotransporter-2 inhibitor, SGLT2i.

**Table 1.**
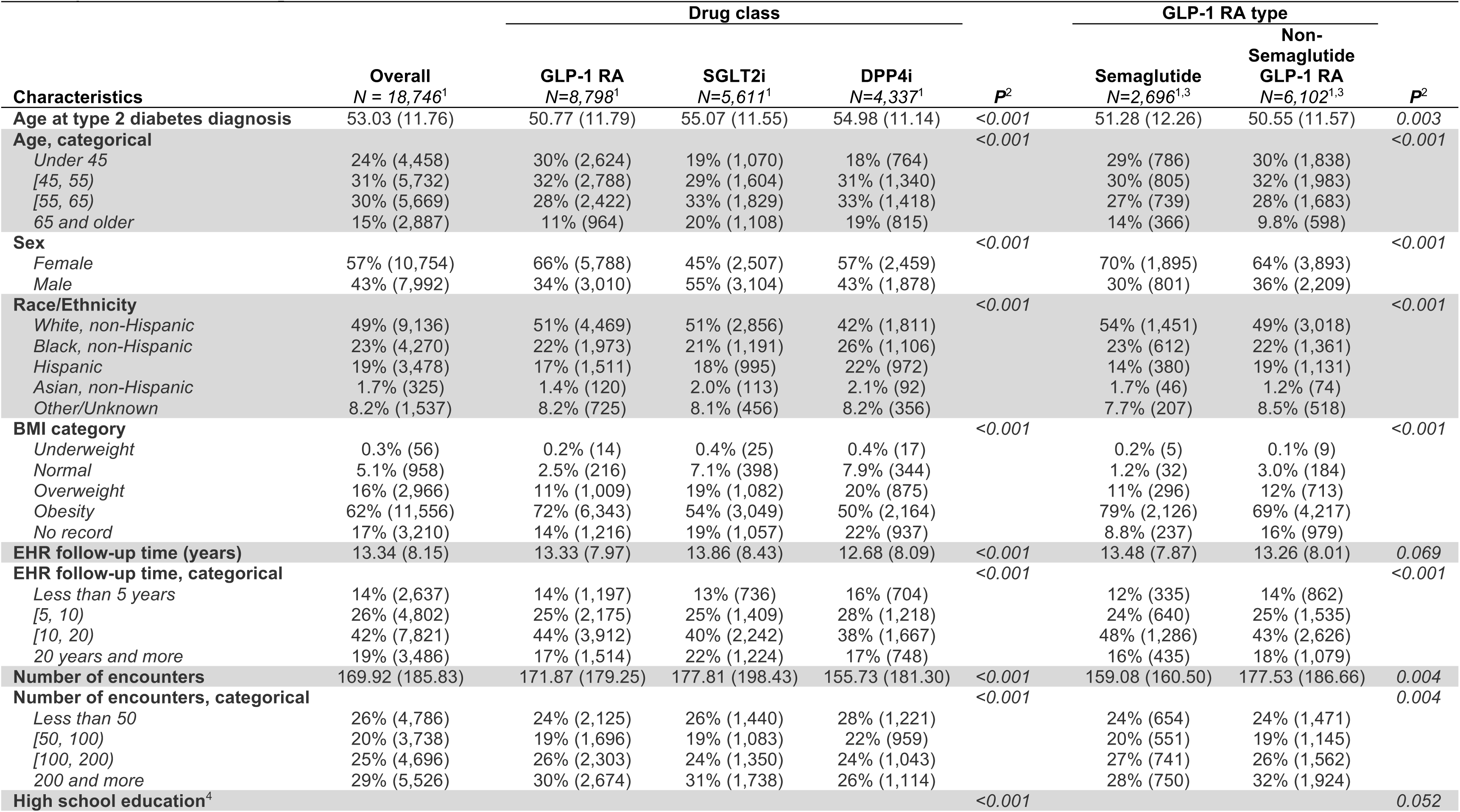
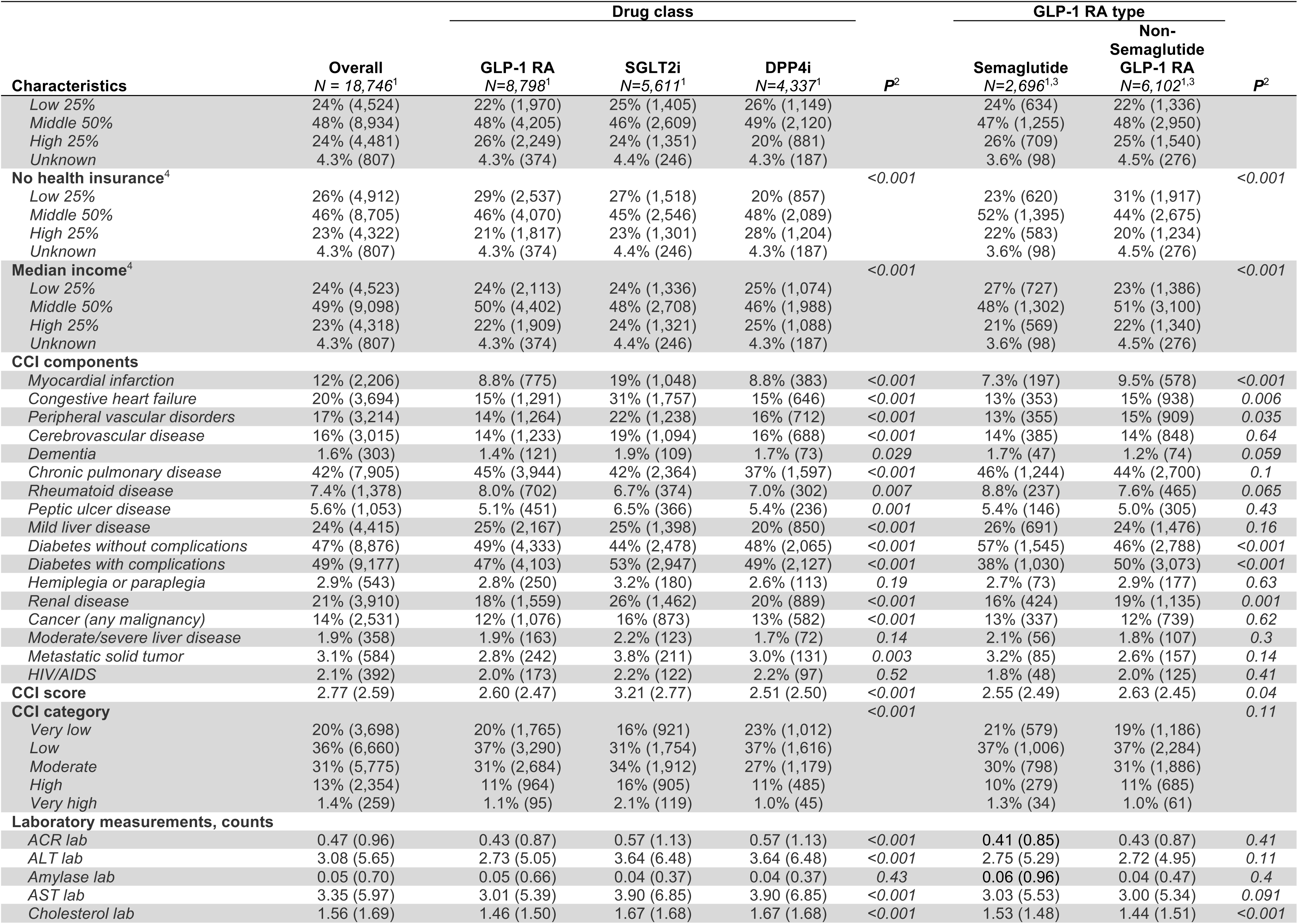
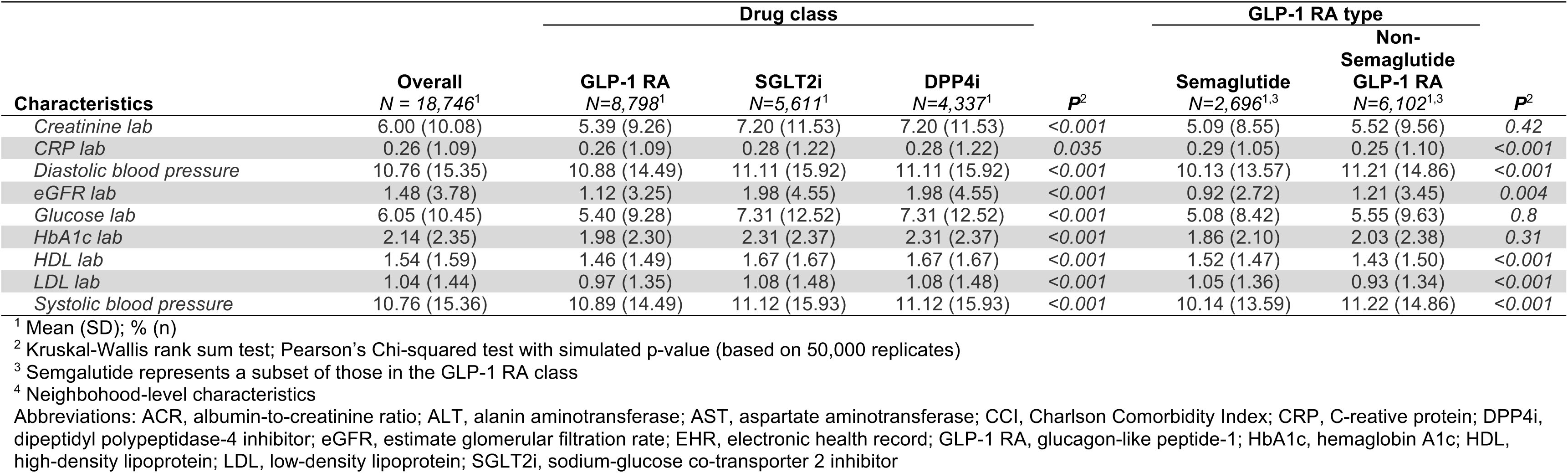
Descriptive characteristics of analytic cohort of individuals with type 2 diabetes in the NIH All of Us Research Program recently prescribed a qualifying anti-diabetes drug on or after January 1, 2018, overall, by drug class, and among those prescribed semaglutide.

Substantial baseline differences were observed across cohorts. The GLP-1 RA cohort was diagnosed with T2D younger (mean age 50.1 years vs. 55.1 for SGLT2i and 55.00 for DPP4i), more likely female (66% vs. 45% for SGLT2i and 57% for DPP4i), and more likely to be obese (72% vs. 54% for SGLT2i and 50% for DPP4i). The SGLT2i cohort, on average, had longer EHR follow-up (13.9 years vs. 13.3 for GLP-1 RA and 12.7 for DPP4i) and had higher CCI scores (3.2 vs 2.6 for GLP-1 RA and 2.5 for DPP4i). Individuals prescribed DPP4i were less likely to be non-Hispanic White (42% vs 51% for GLP-1 RA and SGLT2i).

Patients prescribed semaglutide were considered a subset of the GLP-1 RA cohort. Compared to those prescribed non-semaglutide GLP-1 RAs, those prescribed semaglutide were diagnosed with T2D older (mean age 51.3 vs 50.6), more likely to be female (70% vs. 64%), more likely to be White (54% vs 49%), more likely to be obese (79% vs. 69%), and have lower CCI scores (2.55 vs. 2.63). Despite these differences among individuals prescribed different GLP-1 RAs, those prescribed semaglutide, like the overall GLP-1 RA group, were more likely to be diagnosed with T2D at a younger age, female, and obese compared to those prescribed SGLT2is and DPP4is (Table S1).

### 2.2. Cox proportional hazards PheWAS

#### 2.2.1. GLP-1 RA vs. SGLT2i

In our intention-to-treat PheWAS, there were two phenome-wide significant phenotypes comparing individuals with T2D prescribed GLP-1 RA to those prescribed SGLT2i, after adjusting for multiple testing: inflammatory disease of cervix, vagina, and vulva (hazard ratio (HR) (95% confidence interval (CI)): 0.67 (0.55, 0.81)) and dysthymic disorder (now referred to as persistent depressive disorder; 2.30 (1.53, 3.46)). This PheWAS explored 654 phenotypes with sample sizes ranging from 2,168 for hypertension to 9,196 for “post COVID-19 condition.”

Among the 63 significant (p < Bonferroni-corrected p-value) and suggestive (p < 0.05) phenotypes, GLP-1 RA was associated with a reduced risk for 21% (n=13) and an increased risk for 79% (n=50), relative to individuals prescribed SGLT2i (Figure 2A).

**Figure 2.**
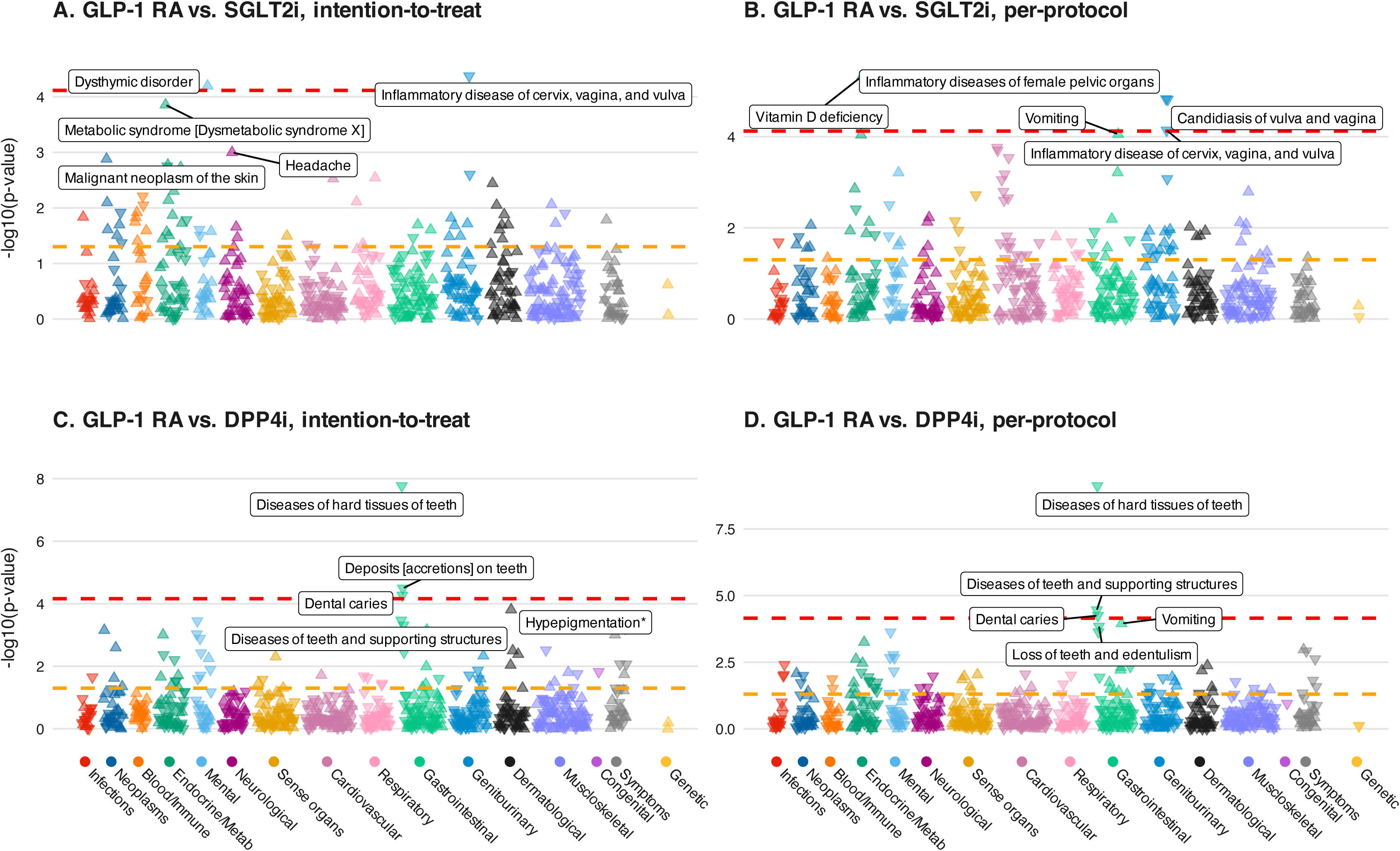
PheWAS plots comparing GLP-1 RA use with SGLT2i (panels A and B) and DPP4i (panels C and D) under intention-to-treat (panels A and C) and per- protocol (panels B and D) approaches. Upward and downward triangles represent RMST difference p-values < 0.05 for traits with increased and decreased time to diagnosis for GLP-1 RA, respectively. Labeled points represent the five largest absolute RMST differences, regardless of p-value. Dashed, horizontal red line indicates phenome-wide significance p-value threshold corrected for multiple testing, while dashed, horizontal orange line represents suggestive p-value threshold of 0.05. P-values and 95% CI are reported in Tables S2 and S3.

GLP-1 RA prescription was most commonly associated with increased risk of endocrine/metabolic disorders (n=11 each). The top 5 suggestive phenotypes (i.e., with smallest p-value) were metabolic syndrome [dysmetabolic syndrome X] (2.05 (1.10, 1.43)), headache (1.25 (1.10, 1.43)), malignant neoplasm of the skin (1.38 (1.13, 1.68)), disorders of parathyroid gland (1.49 (1.16, 1.91), and hyperglycemia (0.83 (0.74, 0.93)).

In our per-protocol PheWAS, there were two phenome-wide significant phenotypes, exploring 667 phenotypes with sample sizes ranging from 2,168 for hypertension to 9,198 for “post COVID-19 condition” (Figure 2B). GLP-1 RA prescription was significantly associated with reduced risk of inflammatory diseases of female pelvic organs (0.58 (0.45, 0.74)), candidiasis of vulva and vagina (0.44 (0.31, 0.64)), and inflammatory disease of cervix, vagina, and vulva (0.60 (0.47, 0.77)) and increased risk of vitamin D deficiency (1.49 (1.26, 1.76)) compared to those prescribed SGLT2is.

Among the 91 significant and suggestive phenotypes, GLP-1 RA was associated with reduced risk for 48% (n=44) and increased risk for 52% (n=47). GLP-1 RA prescription was most commonly associated with reduced risk of cardiovascular diagnoses (n=18) and increased risk of genitourinary diagnoses (n=10).

#### 2.2.2. GLP-1 RA vs. DPP4i

Our intention-to-treat PheWAS explored 733 phenotypes with sample sizes ranging from 1,910 for hypertension to 7,532 for hypertrophic (pulmonary) osteoarthropathy.

There were three phenome-wide significant phenotypes comparing individuals with T2D prescribed GLP-1 RA to those prescribed DPP4i, after adjusting for multiple testing: diseases of hard tissues of teeth (0.54 (0.44, 0.67)), deposits [accretions] on teeth (0.40 (0.26, 0.62)), and dental caries (0.63 (0.50, 0.79)) (Figure 2C).

Among the 67 significant and suggestive phenotypes, GLP-1 RA was associated with reduced risk for 54% (n=36) and increased risk for 46% (n=31), relative to individuals prescribed DPP4i. GLP-1 RA prescription was most commonly associated with reduced risk of gastrointestinal and mental disorders (n=7 each) and increased risk of endocrine/metabolic and dermatological disorders (n=5 each). The top 5 suggestive phenotypes (i.e., with smallest p-value) were hyperpigmentation (1.55 (1.24, 1.94)), diseases of teeth and supporting structures (0.73 (0.62, 0.87), alcohol abuse and dependence (0.56 (0.40, 0.77), pain in limb (0.84 (0.76, 0.93), and loss of teeth and edentulism (0.55 (0.39, 0.77)).

In our per-protocol PheWAS, there were four phenome-wide significant phenotypes, exploring 718 phenotypes with sample sizes ranging from 1,910 for hypertension to 7,532 for hypertrophic (pulmonary) osteoarthropathy. Among these, GLP-1 RA was associated with reduced risk of disease of hard tissues of teeth (0.43 (0.33, 0.56)), diseases of teeth and supporting structures (0.63 (0.51, 0.79)), and dental caries (0.56 (0.42, 0.74)) (Figure 2D).

Among the 83 significant and suggestive phenotypes, GLP-1 RA was associated with reduced risk for 63% (n=52) and increased risk for 37% (n=31). GLP-1 RA prescription was most commonly associated with reduced risk of endocrine/metabolic disorders (n=8) and increased risk of gastrointestinal diagnoses (n=7). The top 5 suggestive phenotypes (i.e., with smallest p-value) were vomiting (1.46 (1.20, 1.76)), loss of teeth and edentulism (0.43 (0.28, 0.66)), deposits [accretions] on teeth 0.38 (0.23, 0.64)), alcohol abuse and dependence (0.46 (0.31, 0.70)), and vitamin D deficiency (1.32 (1.13, 1.55)).

#### 2.2.3. Semaglutide vs. SGLT2i

Our intention-to-treat PheWAS explored 337 phenotypes with sample sizes ranging from 1,104 for hypertension to 4,216 for acute cough. There was one phenome-wide significant phenotype comparing individuals with T2D prescribed semaglutide to those prescribed SGLT2i, after adjusting for multiple testing: vaginitis and vulvovaginitis (0.46 (0.32, 0.68)) (Figure 3A).

**Figure 3.**
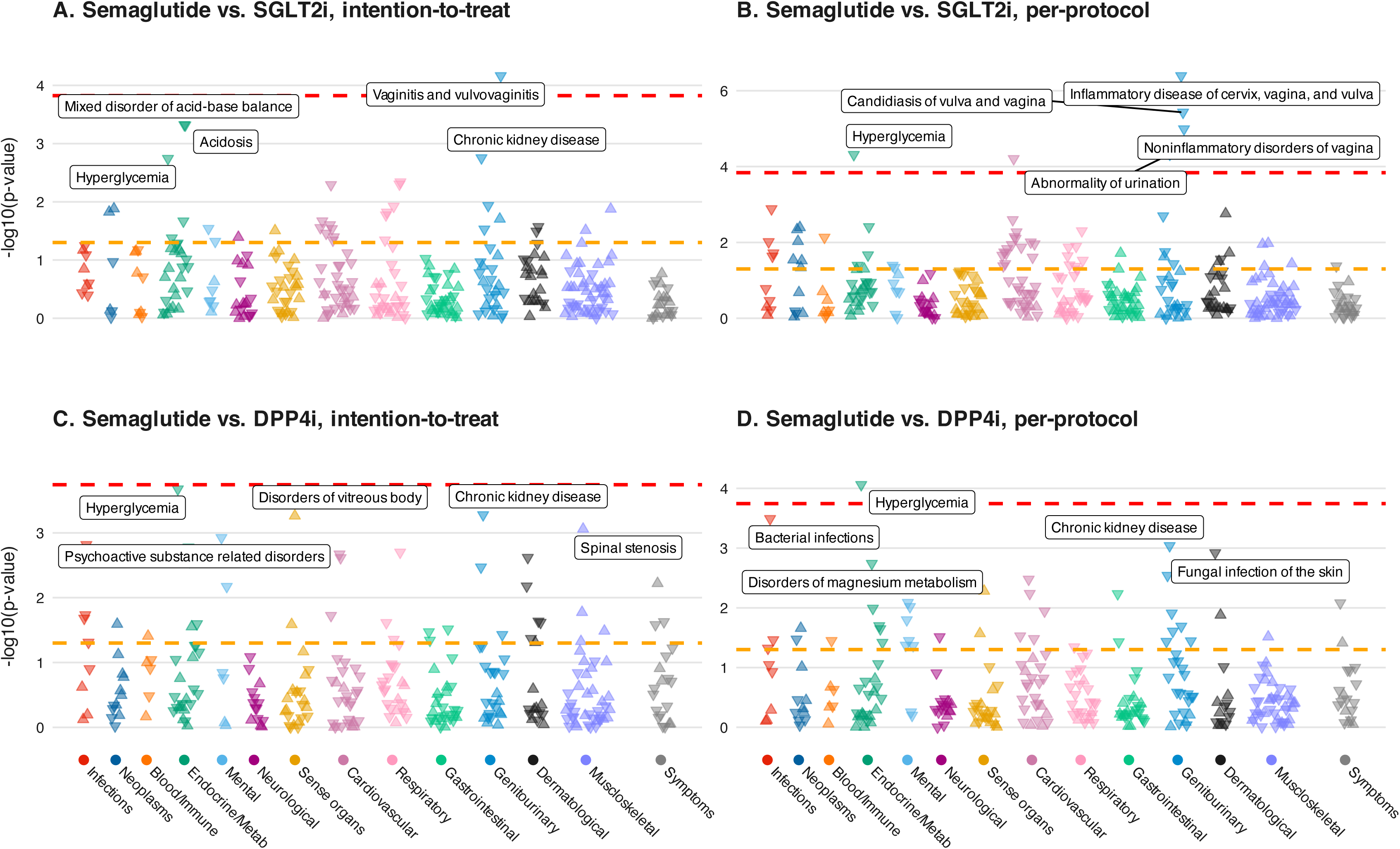
PheWAS plots comparing semaglutide use with SGLT2i (panels A and B) and DPP4i (panels C and D) under intention-to-treat (panels A and C) and per- protocol (panels B and D) approaches. Upward and downward triangles represent RMST difference p-values < 0.05 for traits with increased and decreased time to diagnosis for semaglutide prescription, respectively. Dashed, horizontal red line indicates phenome-wide significance p-value threshold corrected for multiple testing, while dashed, horizontal orange line represents suggestive p-value threshold of 0.05. Labeled points represent the five largest absolute RMST differences, regardless of p-value. P-values and 95% CI are reported in Tables S4 and S5.

Among the 35 significant and suggestive phenotypes, semaglutide was associated with reduced risk for 77% (n=27) and increased risk for 23% (n=8), relative to individuals prescribed SGLT2i. Semaglutide prescription was most commonly associated with reduced risk of cardiovascular diagnoses (n=8) and increased risk of neoplasm diagnoses (n=3). The top 5 suggestive phenotypes (i.e., with smallest p-value) were mixed disorder of acid-base balance (0.46 (0.32, 0.68)), acidosis (0.51 (0.35, 0.74)), chronic kidney disease (0.67 (0.52, 0.86)), hyperglycemia (0.74 (0.61, 0.89)), and melanocytic nevi (1.60 (1.17, 2.20)).

The per-protocol PheWAS explored 349 phenotypes with sample sizes ranging from 1,104 for hypertension to 4,216 for acute cough. Semaglutide prescription was associated with reduced risk for six phenome-wide significant phenotypes: inflammatory disease of cervix, vagina, and vulva (0.33 (0.21, 0.51)), candidiasis of vulva and vagina (0.26 (0.15, 0.46)), noninflammatory disorders of vagina (0.41 (0.27, 0.61)), abnormality of urination (0.62 (0.50, 0.78)), hyperglycemia (0.60 (0.46, .77)), and cardiac arrhythmia and conduction disorders (0.52 (0.38, 0.72)) (Figure 3B).

Among the 60 significant and suggestive phenotypes, semaglutide was associated with reduced risk for 73% (n=44) and increased risk for 27% (n=16). Semaglutide prescription was most commonly associated with reduced risk of cardiovascular diagnoses (n=16) and increased risk of neoplasm diagnoses (n=6). The top 5 suggestive phenotypes (i.e., with smallest p-value) were vaginitis and vulvovaginitis (0.39 (0.24, 0.64)), fungal infections (0.67 (0.52, 0.86)), localized swelling, mass and lump of skin and subcutaneous tissue (1.86 (1.26, 2.75)), chronic kidney disease (0.59 (0.42, 0.82)), cardiomyopathy (0.45 (0.27, 0.76)).

#### 2.2.4. Semaglutide vs. DPP4i

Our intention-to-treat PheWAS explored 278 phenotypes with sample sizes ranging from 698 for hypertension to 2,630 for acidosis. After adjusting for multiple testing, there were no phenome-wide significant phenotypes comparing individuals with T2D prescribed semaglutide to those prescribed DPP4i (Figure 3C).

Among the 40 suggestive phenotypes, semaglutide was associated with reduced risk for 72.5% (n=29) and increased risk for 27.5% (n=11), relative to individuals prescribed DPP4i. Semaglutide prescription was most commonly associated with reduced risk of dermatological diagnoses (n=5) and increased risk of musculoskeletal diagnoses (n=4). The top 5 suggestive phenotypes (i.e., with smallest p-value) were hyperglycemia (0.67 (0.54, 0.83)), chronic kidney disease (0.61 (0.46, 0.81)), disorders of vitreous body (2.18 (1.40, 3.39)), spinal stenosis (1.64 (1.22, 2.19)), and psychoactive substance related disorders (0.54 (0.37, 0.78)).

The per-protocol PheWAS explored 280 phenotypes with sample sizes ranging from 698 for hypertension to 2,640 for altered mental status, unspecified. Semaglutide was significantly associated with reduced risk of hyperglycemia (0.57 (0.43, 0.76)) (Figure 3D).

Among the 42 significant and suggestive phenotypes, semaglutide was associated with reduced risk for 86% (n=36) and increased risk for 114% (n=6). Semaglutide prescription was most commonly associated with reduced risk of endocrine/metabolic and genitourinary diagnoses (n=7 each) and increased risk of sense organ diagnoses (n=2). The top 5 suggestive phenotypes (i.e., with smallest p-value) were bacterial infections (0.52 (0.37, 0.74)), chronic kidney disease (0.53 (0.36, 0.77)), fungal infection of the skin (0.59 (0.43, 0.81)), disorders of magnesium metabolism (0.42 (0.24, 0.72)), and renal failure (0.61 (0.44, 0.84)).

#### 2.2.5. Restricted mean survival time

In phenome-wide comparisons of time-to-diagnosis, GLP-1 RA prescription was associated with heterogeneous RMST differences relative to SGLT2i and DPP4i prescription (Supplementary Section 2). Compared to SGLT2i, GLP-1 RA prescription was significantly associated with delayed diagnosis of inflammatory gynecologic conditions and dysthymic disorder in both intention-to-treat and per-protocol analyses (Figure S1). Across 58 significant and suggestive phenotypes in the intention-to-treat comparison, GLP-1 RA was associated with earlier diagnoses for 74% of traits and later diagnoses for 26%. This pattern was more balanced in per-protocol analyses (53% earlier vs. 47% later), with cardiovascular and genitourinary traits showing the strongest class-specific signals. Comparisons between GLP-1 RA and DPP4i prescriptions revealed significantly later diagnoses for diseases of the teeth and limb pain in intention-to-treat analysis and a comparable distribution of earlier and later diagnoses across phenotypes in both analytic approaches. When analyses were restricted to semaglutide, a stronger tendency toward delayed diagnosis was observed: semaglutide was associated with later time to diagnosis for 78–86% of traits compared to SGLT2i and 79–86% of traits compared to DPP4i, with consistent enrichment in cardiovascular and endocrine/metabolic conditions. Across all analyses, several phenotypes—such as vaginitis and vulvovaginitis, hyperglycemia, chronic kidney disease, and cardiac arrhythmias—exhibited large RMST differences, with magnitudes often exceeding 25 days over three years.

## 3. DISCUSSION

GLP-1 RAs, including semaglutide (e.g., Ozempic, Wegovy), have garnered widespread attention, largely due to media coverage of their substantial weight loss effects in the wake of semaglutide’s FDA approval for weight management in 2021. As many as 1 in 8 Americans have self-reported GLP-1 RA use,^66^ with rapidly growing uptake.^22,23,26^ This study compared diagnoses associated with GLP-1 RA and semaglutide prescription to those with SGLT2 and DPP4 inhibitors between January 2018 and October 2023 among individuals with T2D in the NIH All of Us Research Program.

Our phenome-wide association study identified four key clinically meaningful findings. First, GLP-1 RAs were consistently associated with reduced risk of genitourinary infections in women compared to SGLT2is, with even stronger protective effects seen specifical with semaglutide (PP HR 0.33 [95% CI 0.21, 0.51] for inflammatory disease of cervix, vagina, and vulva). Second, semaglutide demonstrated superior protection against cardiac arrhythmias and conduction disorders compared to SGLT2is (PP HR 0.52 [95% CI 0.38, 0.71]), suggesting potential cardiovascular benefits beyond those previously documented for major adverse cardiac events.^58,67,68^ Third, compared to DPP4is, GLP-1 RAs were significantly associated with reduced risk of dental conditions (PP HR 0.43 [95% CI 0.33, 0.56] for diseases of hard tissues of teeth), a novel finding with implications for oral health management in patients with T2D. Fourth, semaglutide specifically suggested reduced risk of chronic kidney disease compared to both alternative medication classes (PP HR 0.59 [95% CI 0.42, 0.82] versus SGLT2is and HR 0.53 [95% CI 0.36, 0.77] versus DPP4is), supporting its potential renoprotective effects in real-world settings.

Our findings corroborate and extend those reported by Xie and colleagues^49^ in their comprehensive analysis of GLP-1 RA effects. Like their study, we observed reduced risk of genitourinary conditions among GLP-1 RA users compared to SGLT2i users, particularly inflammatory diseases of female pelvic organs (ITT HR 0.73 [95% CI 0.61, 0.88], suggestive in our study, versus ITT HR 0.63 [95% CI 0.56, 0.71] in Xie et al.), aligning with existing literature documenting increased risk of vulvovaginal candidiasis among female SGLT2i users with T2D.^69–71^ However, our analyses reveal more nuanced patterns, especially for semaglutide-specific effects. While Xie found no significant association between GLP-1 RAs and cardiac arrhythmias (HR 1.02 [95% CI 0.99, 1.04]), our semaglutide-specific analysis demonstrated substantial protection against cardiac arrhythmia and conduction disorders (PP HR 0.52 [95% CI 0.38, 0.72] (Table 2)) compared to SGLT2is. Semaglutide has been shown to reduce major cardiovascular events,^58,72,73^ but its comparative effectiveness for arrhythmia outcomes remains understudied.

**Table 2.**
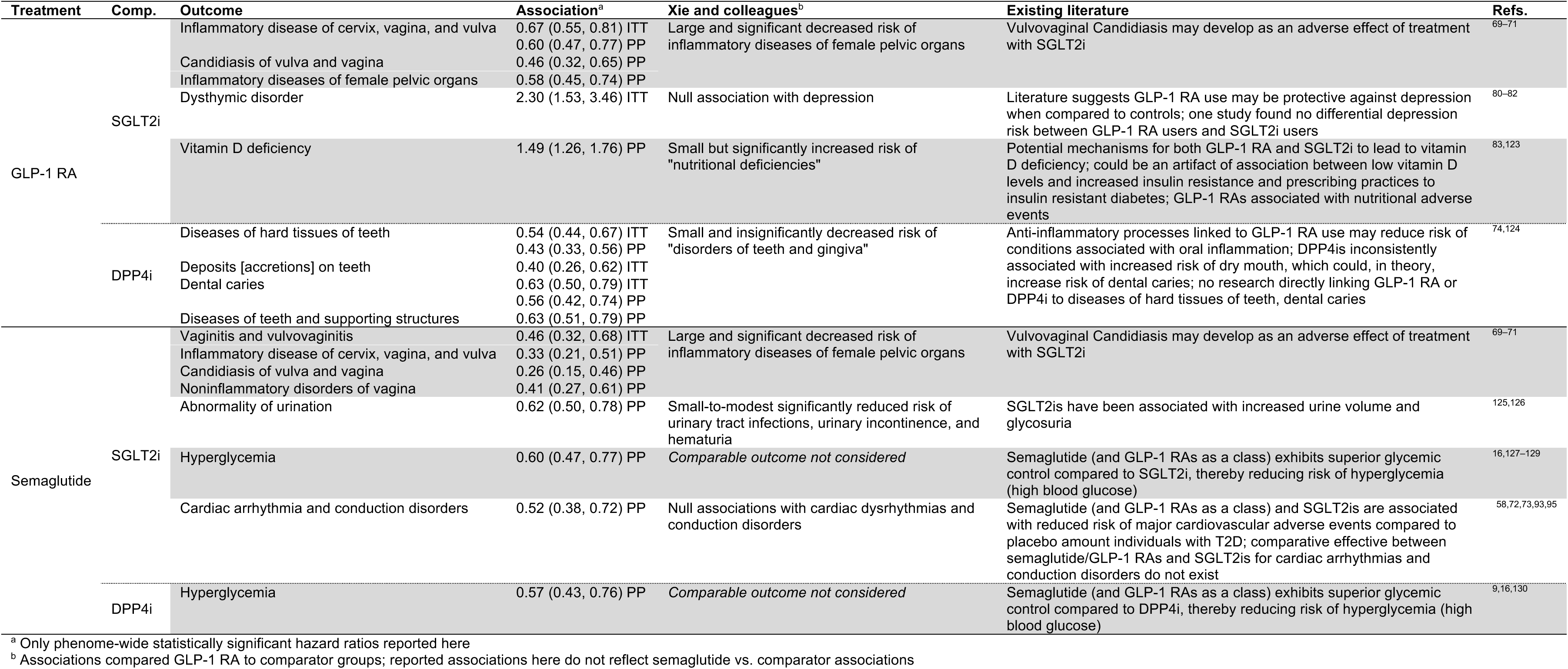
Comparison of statistically significant results in this study with those reported by Xie and colleagues and existing literature.

Similarly, our finding of reduced dental disease risk with GLP-1 RAs differs from Xie’s more modest, non-significant reductions in dental conditions. Anti-inflammatory processes associated with GLP-1 RA action may reduce oral inflammation risk,^74,75^ while limited evidence suggests dry mouth from DPP4i use^76,77^ might facilitate dental caries development.^78,79^ Interestingly, while Xie and colleagues reported reduced substance use and psychotic disorders among GLP-1 RA users, we identified potentially increased risk of dysthymic disorder (ITT HR 2.30 [95% CI 1.53, 3.46]) compared to SGLT2i users. Most existing studies focus on major depressive disorder rather than dysthymia.^80–82^ Xie and colleagues and the existing literature also suggest potential nutritional side effects associated with GLP-1 RAs,^49,83^ consistent with our finding of increased risk of vitamin D deficiency relative to SGLT2i users (PP HR 1.49 [95% CI 1.26, 1.76]).

RMST analyses revealed systematic differences in time to diagnosis, though often modest (typically <1 month over 3 years). However, semaglutide led to longer delays in several phenotypes, including a 78.5-day delay in hyperglycemia diagnosis, highlighting the value of combining relative and absolute measures to assess clinical impact and suggesting that observed patterns may reflect differences in surveillance or disease progression across therapies.

GLP-1 RA PheWAS were better powered (n=8,798) than semaglutide-specific analyses (n=2,696). GLP-1 RA analyses may be affected by heterogeneity in effects^16,29,84–86^ and prescribing practices,^87,88^ as shown by statistically significant baseline covariate differences between semaglutide and other GLP-1 RAs (Table 1).

These findings have important implications for clinical practice and public health oversight. For clinical decision support, providers should consider sex-specific outcomes when selecting second-line therapies, potentially prioritizing GLP-1 RAs, especially semaglutide, for women at risk of genitourinary infections. The observed cardioprotective effects of semaglutide against arrhythmias may warrant consideration in patients with or at risk for cardiac conduction disorders. At the same time, renoprotective benefits could inform medication selection for patients with early kidney dysfunction. For patient education, these data support discussing the potential reduced risk of dental problems with GLP-1 RAs compared to DPP4is, while highlighting monitoring for potentially emerging risks such as dysthymic disorder and vitamin D deficiency. From a regulatory perspective, the differential effects observed between semaglutide and other GLP-1 RAs highlight the need for drug-specific rather than class-wide safety assessments, particularly as telehealth prescribing limits systematic follow-up.

This nuanced approach is essential because GLP-1 RAs vary by duration of action and pharmacodynamic features, which influence prescribing decisions based on glycemic targets, weight loss goals, side effect profiles, and patient preferences. While GLP-1 RAs typically offer the greatest reductions in HbA1c and weight,^9,89–92^ SGLT2is and GLP-1 RAs provide cardiovascular and renal benefits.^93–95^ DPP4is are weight-neutral.^9,96,97^ The emerging pharmacogenomic literature, such as Dawed and colleagues’ identification of *ARRB1* variants with implications for the timing of GLP-1 RA treatment initiation,^85^ and Jakhar and colleagues’ work on inter-individual variation in drug response (primarily focused on exenatide and liraglutide)^86^ further supports the need for prescription in prescribing decisions. However, combination therapy, such as GLP-1 RA combined with SGLT2is, requires additional investigation.^98–100^

This study has several strengths. First, the use of propensity score matching with a small caliper, in theory, reduces systematic baseline differences between treated and untreated participants, mitigating concerns related to confounding bias. Second, we present relative (via hazard ratios) and absolute (via RMST differences) differences in risk, better contextualizing results, and enhancing interpretability. Third, AOU is a large, diverse cohort that spans the USA, which can capture differences and trends potentially missed or absent using single medical center-based or regional cohorts. Fourth, we explore semaglutide-specific analyses. While many recent analyses consider GLP-1 RAs as a class, in part to increase sample size and because of relatively similar pharmacogenomic actions of drugs within the class, there may be heterogeneity in how different GLP-1 RAs are prescribed. Additionally, semaglutide has received targeted attention recently, so results looking at semaglutide specifically are particularly relevant.

This study also has several important limitations. First, as with any agnostic association analysis using observational data, positive results require follow-up mechanism-focused studies, and despite efforts to reduce measured confounding, unmeasured confounding may still be present. Second, propensity score models are sensitive to model specification,^101^ which may be misspecified here. Additional analyses can be considered, exploring various propensity models (e.g., higher dimensional, wider caliper width), including non-parametric and data-driven approaches. Third, our focus on T2D patients limits the generalizability to the rapidly growing population using GLP-1 RAs for weight management (prescriptions rose 27% for GLP-1 RAs and 41% for semaglutide between March and June 2024).^26^ However, sample size constraints prevented comparisons with other anti-obesity medications. Fourth, GLP-1 RA use is likely underreported in EHR data, particularly for telehealth prescriptions - only 5.3% of All of Us participants had documented GLP-1 RA prescriptions versus 12.5% self-reported use in recent surveys,^66^ potentially biasing our estimates toward the null. More advanced methods to account for underreported exposure status can be explored and incorporated in future analyses.^102^ Finally, prescription records do not guarantee medication adherence, a limitation inherent to all EHR-based pharmacoepidemiology studies.

This study examines downstream impacts of GLP-1 RA prescription compared to two other, common second-line T2D therapies - SGLT2 inhibitors and DPP4 inhibitors - in a national, diverse, real-world cohort of individuals with T2D. We observed that GLP-1 RAs, particularly semaglutide, were associated with reduced risk of several clinically relevant outcomes, including genitourinary infections in women, dental conditions, cardiac arrhythmias, hyperglycemia, and chronic kidney disease. Conversely, we also identified potential adverse associations, such as increased risks of dysthymic disorder and vitamin D deficiency, underscoring the importance of comprehensive safety and effectiveness profiling.

While this study provides an initial view into the broader health implications of GLP-1 RA prescription as a time of high public interest and rapidly growing uptake, more work is needed. Future work should investigate the effects of GLP-1 RAs in populations without T2D, particularly those using these medications for weight management, and compare outcomes with other anti-obesity therapies. Stratified analyses by age, race/ethnicity, and BMI category in cohorts with sufficient power will be essential to identify population-specific benefits and risks. In parallel, EHR-linked biobanks offer a unique opportunity to explore genetic predictors of individual response to GLP-1 RAs and semaglutide, enabling more personalized treatment strategies.

These findings reinforce the need to consider comorbidities, sex-specific risks, and pharmacogenomic variation in clinical decision-making and point toward the promise of precision prescribing in T2D and obesity care.

## 4. METHODS

### 4.1. Data sources

AOU is a nationally recruited EHR-linked biobank at the US National Institutes of Health, oversampling groups historically underrepresented in biomedical research.^65,103^ Adults can join directly or at over 340 recruitment sites across the US, where they can consent to EHR data access, provide biospecimens, and complete surveys.

We used the curated data repository version 8^104^ and incorporated demographic (n=633,547), International Classification of Disease (ICD; 9^th^ and 10^th^ revisions) code (n=344,321), and medication data (n=334,162). ICD codes were mapped to broader yet clinically meaningful phenotypes called phecodes (version X,^105^ described elsewhere^106^; see Section 4.3).

The analytic cohort represents individuals with self-reported male or female sex assigned at birth and a T2D phecode [EM_202.2; underlying ICD codes shown in Table S1] code who received one of the qualifying medications (including GLP-1 RA, SGLT2i, or DPP4i). The cohort was further restricted to individuals who (i) were prescribed a qualifying drug after January 1, 2018, after their T2D diagnosis, (ii) had at least one year of medical records prior to their first qualifying drug prescription (*index date*) (iii) had no qualifying drug prescription within 120 days of the index date, (iv) were not prescribed multiple qualifying drugs on the index date, and (v) did not have a qualifying contraindication (multiple endocrine neoplasia type II, gastroparesis, low eGFR (<30 mL/min/1.73m^2^), medullary thyroid cancer, hypoglycemia with coma, dialysis, or kidney transplant) in the five years prior to the index date (Table S6). The resulting cohort comprised 18,746 individuals classified into GLP-1 RA (n=8,798), SGLT2i (n=5,611), and DPP4i (n=4,337) initiators focused on the time period from January 1, 2018 (shortly after semaglutide received FDA approval), to October 1, 2023 (administrative cutoff of dataset).

### 4.2. Treatment and comparison groups

Our primary treatments of interest were (i) GLP-1 RAs as a class and (ii) semaglutide as a drug. GLP-1 RAs consisted of albiglutide, beinaglutide, dulaglutide, exenatide, liraglutide, lixisenatide, semaglutide, and tirzepatide (which, despite being a dual GLP-1 and gastric inhibitory polypeptide (GIP) agonist, has been included in GLP-1 RA analyses). Our comparison groups were (i) SGLT2is and (ii) DPP4i. SGLT2is consisted of bexagliflozin, canagliflozin, dapagliflozin, empagliflozin, ertugliflozin, ipragliflozin, luesogliflozin, and sotagliflozin. DPP4i consisted of alogliptin, gemigliptin, linagliptin, saxagliptin, sitagliptin, teneligliptin, and vildagliptin.

Drugs were initially queried using Observational Medical Outcomes Partnership (OMOP) concept IDs associated with each drug name in the All of Us cohort builder.

Because of the roll-up structure present in OMOP, drugs sometimes co-prescribed were obtained (e.g., statins with SGLT2is). Drugs were further filtered such that the name associated with the OMOP concept ID contained one of the drug names listed above.

The resulting set of codes used is presented in Table S7.

### 4.3. Outcomes of interest

Phecodes were first defined in 2010 to facilitate high-throughput genetic analyses (single nucleotide polymorphism-phenotype PheWAS) by aggregating ICD codes into broader yet clinically meaningful phenotypes.^106^ The most recent ICD-to-phecode mapping tables, Phecode X,^105,107^ were designed with ICD-10-CM in mind and define 3,612 phenotypes across 18 groups, ranging from 106 phecodes in the Blood/Immune group to 325 in the Congenital group. Phenotypes defined using ICD code-derived phecodes (version X), mapped using the PheWAS R package (version 0.99.6-1),^108,109^ serve as the outcomes of interest.

For a given treatment-comparator PheWAS, the set of outcomes considered represents all phecodes that occurred in at least 100 individuals in the phenome after the index date (up to 1,026 phecodes in our analyses). Individuals with a record of the outcome within 2 years before the index date were excluded from analyses for that outcome.

We excluded phecodes for obesity, sleep apnea, and diabetes-related traits from the main text, given their complex relationships with coding practices and treatment indications. These are discussed in Supplementary Section 2.

### 4.4. Covariates

Demographic, healthcare utilization, health history, and laboratory covariates were used during the matching portion of our analyses. Demographic factors included age at T2D diagnosis, sex assigned at birth, race/ethnicity (non-Hispanic Asian, non-Hispanic Black, Hispanic, non-Hispanic White, Other/Unknown), last body mass index (BMI) category prior to or on index date, the index date, and neighborhood-level characteristics (proportion with high school education, proportion without health insurance, and median income; 2017 American Community Survey estimates).

Neighborhood-level variables were categorized into four percentile-based groups: low (bottom 25%), middle (middle 50%), high (top 25%), and unknown. Healthcare utilization measures included length of follow-up (years between first ICD code diagnosis and index date) and number of encounters (unique diagnosis days in ICD code data) prior to and including the index date. Health history (any time prior to or on index date) was summarized as indicator variables for the presence of ICD-9-CM and ICD-10-CM codes for each of the 17 components of the Charlson Comorbidity Index^110,111^ (components and corresponding qualifying ICD codes presented in Table S8). These components were mapped using the comorbidity R package (version 1.1.0.9000).^112,113^ Laboratory measurements (Table S9) were included as the *number* of each of the following labs recorded in the two years prior to the index date: aspartate aminotransferase (AST), blood pressure, C-reactive protein, estimated glomerular filtration rate (eGFR), glucose, and hemoglobin A1c (HbA1c). These lab tests were identified by their OMOP concept ID obtained in the AOU cohort builder. Individuals without a record were coded as 0.

### 4.5. Statistical analysis

For each treatment-comparator pair, Cox proportional hazard and restricted mean survival time (RMST) PheWAS. For each outcome (n≥100 after index date), individuals who had a record of the outcome phecode in the two years prior to the index date were removed. Among those who remained, propensity score matching was performed using covariates described above with a caliper width of 0.1. If there were fewer than 50 occurrences of the outcome after matching, no model was fit (up to 766 phecodes met this threshold).

After matching, covariate balance was assessed using standardized mean differences (SMDs), where an SMD < 0.1 indicated adequate balance.^114^ If all covariates exhibited SMD < 0.1, an unadjusted Cox proportional hazards model was fit:

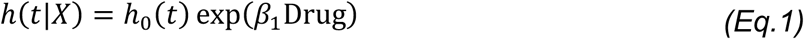

where *h*(*t*|*X*)is the hazard function at time *t*, *h*_0_(*t*) is the baseline hazard, and Drug is an indicator variable for treatment assignment (GLP-1 RA/semaglutide vs. SGLT2i/DPP4i).

If residual imbalance was observed (SMD≥0.1 for any covariate), the model was adjusted for the imbalanced covariates:

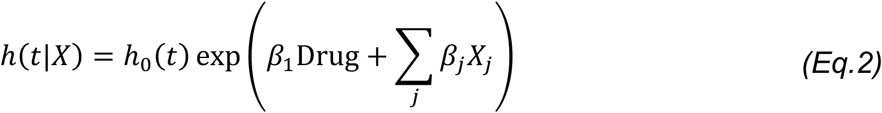

where *X_j_* represents the covariates with SMD≥0.1 after matching.

The proportional hazards assumption was assessed using a Schoenfeld residual test,^115^ and only models with a p-value > 0.05 were retained (up to 736 phecodes met this threshold). Hazard ratios (HRs) and 95% confidence intervals were reported. If there were convergence or separation issues, a Firth-corrected Cox proportional hazards regression model was fit instead.

When interpreting HRs, significant (Bonferroni-corrected p-value; 0.05 / number of models fit) and suggestive (0.05) p-value thresholds were applied. PheWAS, following intention-to-treat and per-protocol approaches, were fit. Under the per-protocol approach, censoring events included discontinuation (no prescription of the index drug recorded within 90 days) and switching (prescription of the comparator drug).

We also estimated restricted mean survival time (RMST) differences between treatment and comparator groups for each outcome that met the proportional hazard assumption. RMST details are presented in Supplementary Section 1.

### 4.6. Software and tools

Analyses were conducted using R 4.4.0. Relevant R packages included coxphf (version 1.13.4)^116–118^ for Firth-corrected Cox proportional hazards models, MatchIt (version 4.7.0)^119,120^ for propensity score matching, and survival (version 3.8-3)^121,122^ for Cox proportional hazards models. The code to carry out the analyses performed in this paper is available on GitHub: https://github.com/RitchieLab/glp1-phewas/.

### 4.7. Ethical considerations

This study was performed using de-identified, publicly available data. Data privacy and reporting requirements defined by the All of Us research program for data use were adhered to.

## Data Availability

NIH All of Us Research Program data is available to individuals at institutions who have a Data Use and Registration Agreement in place and who complete the necessary introductory, regulatory, and access protocols, which are introduced at: https://www.researchallofus.org/register/. The code used to perform analyses conducted in this manuscript is available via GitHub: https://github.com/RitchieLab/glp1-phewas/.

https://researchallofus.org/

## ACKNOWLEDGMENTS

The All of Us Research Program is supported by the National Institutes of Health, Office of the Director: Regional Medical Centers: 1 OT2 OD026549; 1 OT2 OD026554; 1 OT2 OD026557; 1 OT2 OD026556; 1 OT2 OD026550; 1 OT2 OD 026552; 1 OT2 OD026553; 1 OT2 OD026548; 1 OT2 OD026551; 1 OT2 OD026555; IAA #: AOD 16037; Federally Qualified Health Centers: HHSN 263201600085U; Data and Research Center: 5 U2C OD023196; Biobank: 1 U24 OD023121; The Participant Center: U24 OD023176; Participant Technology Systems Center: 1 U24 OD023163; Communications and Engagement: 3 OT2 OD023205; 3 OT2 OD023206; and Community Partners: 1 OT2 OD025277; 3 OT2 OD025315; 1 OT2 OD025337; 1 OT2 OD025276. In addition, the All of Us Research Program would not be possible without the partnership of its participants.

We gratefully acknowledge All of Us participants for their contributions, without whom this research would not have been possible. We also thank the National Institutes of Health’s All of Us Research Program for making the participant data examined in this study available.

