## Supplementary Materials for "Real-world comparative outcomes of GLP-1 RA and semaglutide prescription among individuals with type 2 diabetes"

### Supplementary Section 1. Restricted mean survival time analyses

1. Methods

We additionally estimated restricted mean survival time (RMST) to compare absolute survival differences. RMST represents the area under the survival curve up to a pre-specified time horizon $\tau$, defined as:

$$\mathrm{RMST}(\tau) = \int_0^{\tau} S(t)dt \qquad \textit{(Eq.S1)}$$

where $S(t)$ is the survival function. The difference in RMST ($\Delta\mathrm{RMST}$) between treatment and comparator drugs was calculated using a 95% confidence interval estimated using a standard error-based method. Unlike the Cox proportional hazards models, RMST estimates were not adjusted for residual covariate imbalance after matching. RMST analyses were conducted at 1-, 2-, and 3-year time horizons (i.e., $\tau$). RMST difference estimates were calculated using the rmst2() function from the survRM2 R package (version 1.0-4)[1,2].

2. Results

2.1. GLP-1 RA vs. SGLT2i

In our intention-to-treat analysis, GLP-1 RA prescription was significantly associated with differences in time to diagnosis at 3 years for inflammatory disease of cervix, vagina, and vulva (difference in days (95% CI): 17.7 (10.1, 25.2)) and dysthymic disorder (-7.4 (-10.9, -3.9)) (Figure S1A).

Of 58 significant and suggestive traits, GLP-1 RA prescription was associated with increased time to diagnosis for 26% (n=15) of traits, and decreased time to diagnosis for 74% (n=43) of traits. Regarding trait categories, GLP-1 RA was most commonly associated with increased times to diagnosis for cardiovascular conditions (n=), and decreased time to diagnosis for dermatological conditions (n=8) compared to SGLT2i prescription. The top 5 suggestive phenotypes (i.e., with smallest p-value) were metabolic syndrome [dysmetabolic syndrome X] (-7.6 (-11.5, -3.5)), vaginitis and vulvovaginitis (10.5 (4.8, 16.2)), headache (-17.1 (-28.5, -5.7)), hyperglycemia (27.8 (9.2, 46.4)), and malignant neoplasm of the skin (-10.0 (-16.7, -3.3)).

In our per-protocol analysis, GLP-1 RA prescription was significantly associated with longer time to diagnosis at 3 years for four phecodes: inflammatory disease of female pelvic organs (21.8 (12.1, 31.5)), inflammatory disease of cervix, vagina, and vulva (21.1 (11.5, 30.8)), candidiasis of vulva and vagina (15.0 (8.0, 22.0)), and rheumatic fever and chronic rheumatic heart diseases (15.3 (7.8, 22.7)).

Of 88 significant and suggestive traits, GLP-1 RA prescription was associated with increased time to diagnosis for 47% (n=41) of traits and reduced time to diagnosis for

53% (n=47) of traits. Regarding trait categories, GLP-1 RA was most commonly associated with increased times to diagnosis for cardiovascular conditions (n=20) and decreased time to diagnosis for genitourinary conditions (n=11) compared to SGLT2i prescription. The top 5 suggestive phenotypes (i.e., with smallest p-value) were vitamin D deficiency (-32.2 (-48.2, -16.1)), rheumatic heart disease (14.2 (6.8, 21.6)), vomiting (-22.6 (-34.7, -10.6)), dysthymic disorder (-7.8 (-12.0, -3.5)), and vitamin deficiencies (-32.0 (-49.6, -14.3)).

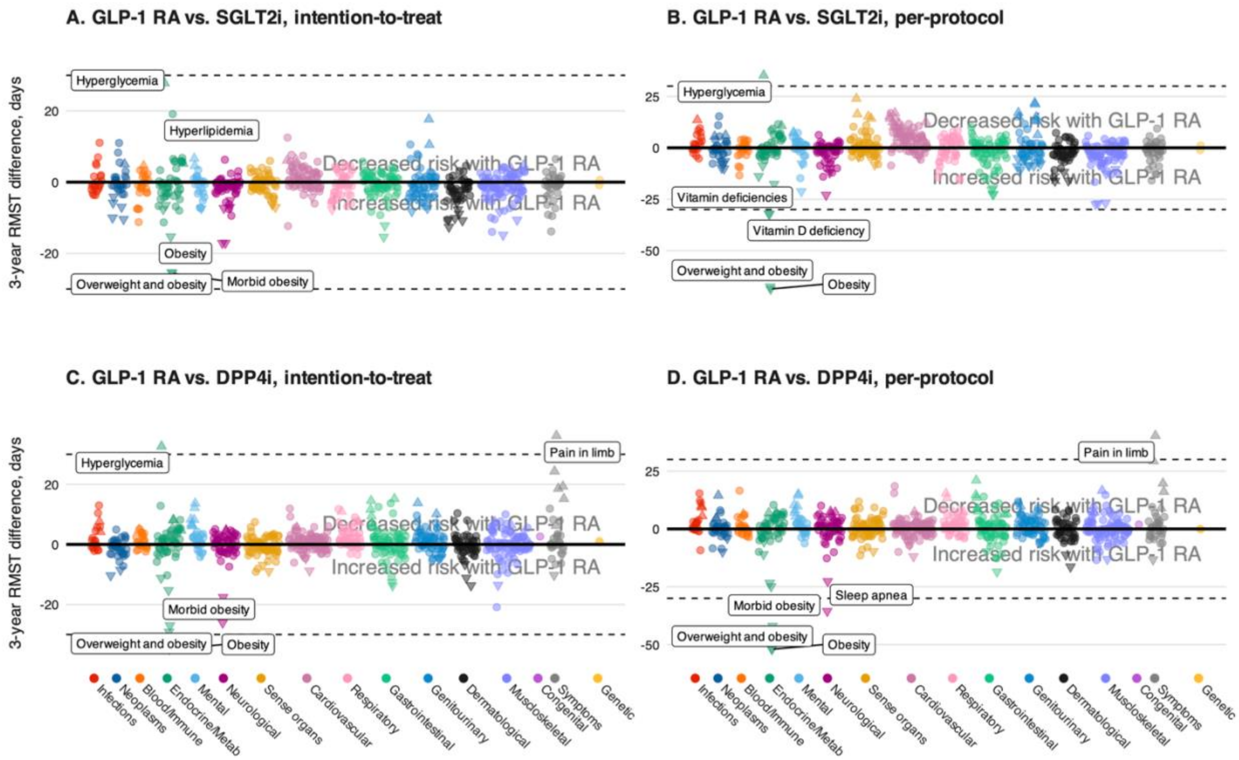


Figure S1. Restricted mean survival time (RMST) plots comparing GLP-1 RA prescription with SGLT2i (panels A and B) and DPP4i (panels C and D) under intention-to-treat (panels A and C) and per-protocol (panels B and D) approaches. Upward and downward triangles represent RMST difference p-values < 0.05 for traits with increased and decreased time to diagnosis for GLP-1 RA, respectively. Labeled points represent the five largest absolute RMST differences, regardless of p-value. P-values and 95% CI are reported in Supplementary Tables 6 and 7.

#### 2.2. GLP-1 RA vs. DPP4i

In our intention-to-treat analysis, there were two statistically significant phenotypes associated with increased time-to-diagnosis comparing GLP-1 RA prescription to DPP4i prescription: diseases of hard tissues of teeth (14.6 (7.6, 21.5)) and pain in limb (36.2 (18.5, 53.9)) (Figure S1C). Of 64 significant and suggestive phenotypes, GLP-1 RA prescription was associated with increased time to diagnosis for 67% (n=43) and decreased time to diagnosis for 33% (n=21). Regarding phenotype categories, GLP-1 RA was most commonly associated with increased times to diagnosis for gastrointestinal conditions (n=8) and decreased time to diagnosis for cardiovascular, dermatological, and genitourinary conditions (n=3 each) compared to

DPP4i prescription. The top 5 suggestive phenotypes (i.e., with smallest p-value) were deposits [accretions] on teeth (7.0 (3.2, 10.8)), allergy (19.3, 8.0, 30.7)), other follicular disorders (-13.7 (-21.8, -5.6)), flat foot [pes planus] (7.5 (3.0, 12.1)), and current tobacco use and nicotine dependence (13.7 (5.2, 22.2)).

In our per-protocol analysis, GLP-1 RA prescription was associated with significantly shorter time to diagnosis at 3 years for diseases of hard tissues of teeth (21.0 (12.1, 29.9)) (Figure S1D). Of 64 significant and suggestive traits, GLP-1 RA prescription was associated with longer time to diagnosis for 60% (n=42) and shorter time to diagnosis for 40% (n=28). Regarding trait categories, GLP-1 RA prescription was most protective against (n=6) and increased risk for (n=4) gastrointestinal conditions compared to DPP4i prescription. The top 5 suggestive phenotypes (i.e., with smallest p-value) were pain in limb (40.3 (18.6, 61.9)), deposits [accretions] on teeth (8.7 (3.8, 13.6)), loss of teeth and edentulism (8.8 (3.5, 14.0)), dental caries (13.5 (5.2, 21.9)), and other follicular disorders (-16.3 (-26.5, -6.2)).

#### 2.3. Semaglutide vs. SGLT2i

In our intention-to-treat analysis, semaglutide prescription was associated with significantly longer time to diagnosis at 3 years for vaginitis and vulvovaginitis (21.6 (11.9, 31.3)) (Figure S2A). Of 36 significant and suggestive traits, semaglutide prescription was associated with increased time to diagnosis for 78% (n=28) of traits and reduced time to diagnosis for 22% (n=8) of traits. Regarding phenotype categories, semaglutide prescription was most commonly associated with increased times to diagnosis for cardiovascular conditions (n=8) and decreased time to diagnosis for neoplasms (n=3) compared to SGLT2i prescription. The top 5 suggestive phenotypes (i.e., with smallest p-value) were acidosis (16.8 (7.4, 26.2)), mixed disorder of acid-base balance (16.0 (6.6, 25.4)), hyperglycemia (41.0 (15.0, 67.0)), chronic kidney disease (23.2 (8.3, 38.2)), and benign neoplasm of the skin (-18.7 (-31.4, -5.9)).

In our per-protocol analysis, semaglutide prescription was associated with significantly longer time to diagnosis at 3 years for six phenotypes: inflammatory disease of cervix, vagina, and vulva (40.8 (25.0, 56.5)), candidiasis of vulva and vagina (29.2 (16.9, 41.6)), hyperglycemia (78.5 (43.2, 113.8)), noninflammatory disorders of vagina (38.8 (20.6, 56.9)), abnormality of urination (66.9 (34.6, 99.2)), and vaginitis and vulvovaginitis (25.6 (12.4, 38.7)) (Figure S2A). Of 51 significant and suggestive traits, semaglutide prescription was associated with increased time to diagnosis for 78% (n=40) of traits and reduced time to diagnosis for 22% (n=11) of traits. Regarding phenotype categories, semaglutide prescription was most commonly associated with increased times to diagnosis for cardiovascular conditions (n=14) and decreased time to diagnosis for neoplasms (n=5) compared to SGLT2i prescription. The top 5 suggestive phenotypes (i.e., with smallest p-value) were cardiac arrhythmia and conduction disorders (40.6 (18.5, 62.7)), hemangioma and lymphangioma (-19.0 (-31.5, -6.5)), localized swelling, mass and lump of skin and subcutaneous tissue (-25.0 (-41.7, -8.3)), paroxysmal atrial fibrillation (17.5 (5.6, 29.3)), and chronic kidney disease (28.4 (8.3, 48.4)).

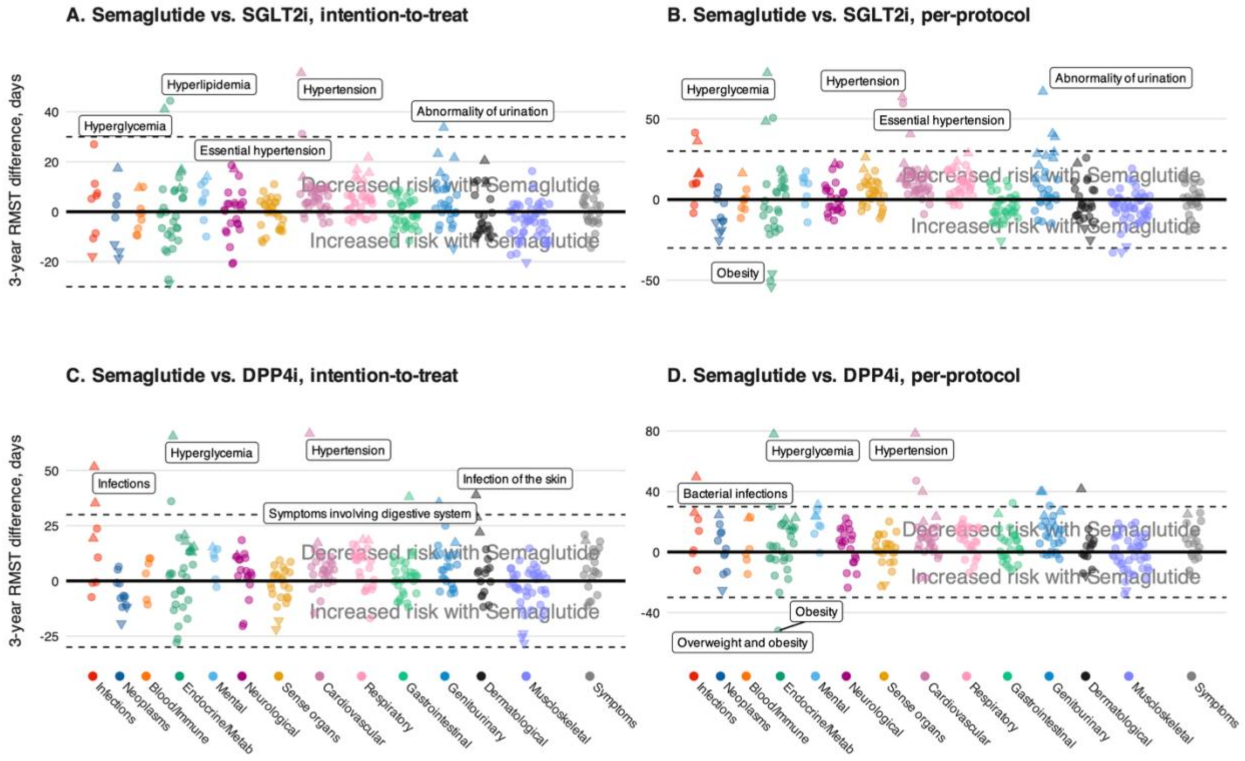


Figure S2. Restricted mean survival time (RMST) plots comparing semaglutide prescription with SGLT2i (panels A and B) and DPP4i (panels C and D) under intention-to-treat (panels A and C) and per-protocol (panels B and D) approaches. Upward and downward triangles represent RMST difference p-values < 0.05 for traits with increased and decreased time to diagnosis for semaglutide prescription, respectively. Labeled points represent the five largest absolute RMST differences, regardless of p-value. P-values and 95% CI are reported in Supplementary Tables 8 and 9.

#### 2.4. Semaglutide vs. DPP4i

In our intention-to-treat analysis, semaglutide prescription was associated with significantly longer time to diagnosis at 3 years for hyperglycemia (65.5 (32.4, 98.6)) (Figure S2C). Of 34 significant and suggestive traits, GLP-1 RA prescription was associated with increased time to diagnosis of 79% (n=27) of traits and reduced time to diagnosis for 21% (n=7) of traits. Regarding trait categories, semaglutide use was most protective against endocrine/metabolic conditions (n=7) and increased risk for musculoskeletal conditions (n=3) compared to DPP4i prescription. The top 5 suggestive phenotypes (i.e., with smallest p-value) were chronic kidney disease (35.5 (16.8, 54.1)), bacterial infections (35.2 (16.6, 53.9)), disorder of vitreous body (-18.0 (-28.5, -7.4)), disorders of magnesium metabolism (20.7 (8.4, 33.0)), and spinal stenosis (-28.1 (-44.8, -11.4)).

In our per-protocol analysis, semaglutide prescription was not associated with differential time to diagnosis at 3 years compared to DPP4i (Figure S2D). Of 29 significant and suggestive traits, semaglutide was associated with increased time to diagnosis for 86% (n=25) of traits and reduced time to diagnosis for 14% (n=4) of traits. Regarding trait categories, semaglutide use was most protective against genitourinary and endocrine/metabolic conditions (n=5 each). Semaglutide use demonstrated increased risk for one phecode in neoplasms, dermatological, musculoskeletal, and sense organ categories. The top 5 suggestive phenotypes (i.e., with smallest p-value) were bacterial infections (49.5 (22.8, 76.2)), hyperglycemia (77.8 (35.0, 120.6)), chronic kidney disease (39.8 (15.1, 64.5)), cardiac arrhythmia and conduction disorders (39.9 (13.5, 66.3)), and fungal infections of the skin (41.6 (13.5, 69.8)).

### Supplementary Section 1. On obesity, diabetes, and sleep disorders

GLP-1 RAs, based on their established pharmacologic mechanisms[3–7] and a wealth of clinical trial and real-world evidence,[8–12] are unequivocally linked to weight loss. However, our preliminary analyses revealed positive associations between GLP-1 RA use and *ICD code-derived phecodes* for overweight and obesity (EM_236 and its child code), (pre)diabetes (EM_202 and its child code; EM_204.5), and sleep disorders (NS_333 and its child codes, including NS_333.1, sleep apnea). The plots corresponding to preliminary analyses, where the results for these phecodes were retained, are shown in Figure S*3*. We examined these contradictory findings and determined that they are false positive results—artifacts of clinical coding practices amid the increase and shortages of GLP-1 RA prescriptions.

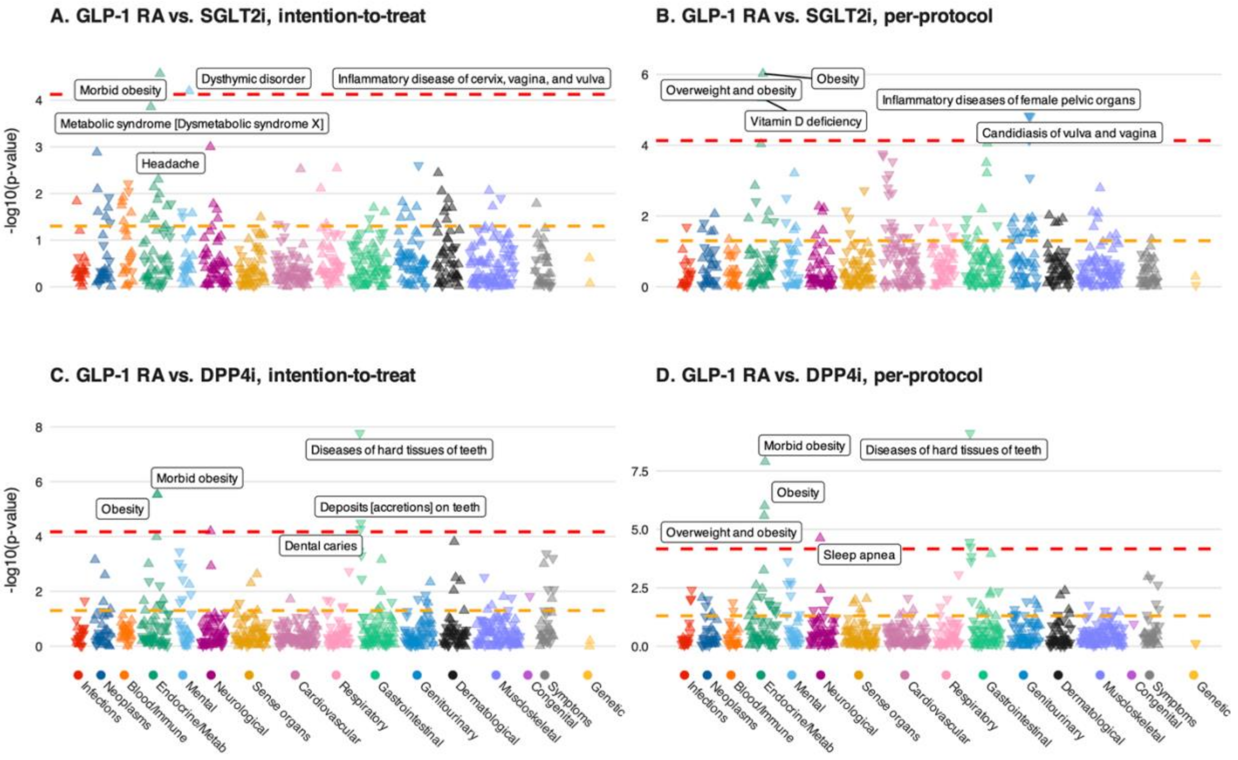


Figure S3. PheWAS plots comparing GLP-1 RA use with SGLT2i (panels A and B) and DPP4i (panels C and D) under intention-to-treat (panels A and C) and per-protocol (panels B and D) approaches. Unlike Figure 2, it includes phecodes for overweight and obesity, (pre)diabetes, and sleep disorders (including sleep apnea). Upward and downward triangles represent RMST difference p-values < 0.05 for traits with increased and decreased time to diagnosis for GLP-1 RA, respectively. Labeled points represent the five largest absolute RMST differences, regardless of p-value. The dashed horizontal red line indicates a phenome-wide significance p-value threshold corrected for multiple tests. In contrast, the dashed horizontal orange line represents a suggestive p-value threshold of 0.05. P-values and 95% CI are reported in Tables S2 and S3.

We compared BMI trajectories with the timing of obesity phecodes and found no association. Individuals with BMI ≥ 30 had BMIs that qualified them for obesity-related ICD codes; however, these codes were recorded much later or, in some cases, were never recorded. Figure S4 shows a sample of 9 individuals diagnosed with morbid obesity after their GLP-1 RA initiation who had at least 3 BMI measurements within a year of treatment initiation. We see that the timing of a BMI measurement ≥ 40 is not correlated with the timing of a morbid obesity diagnosis. There are individuals whose BMI is below the threshold (e.g., individual in gray) at the time of diagnosis and individuals who had qualifying BMI measurements well before the diagnosis (e.g., individuals in black and orange).

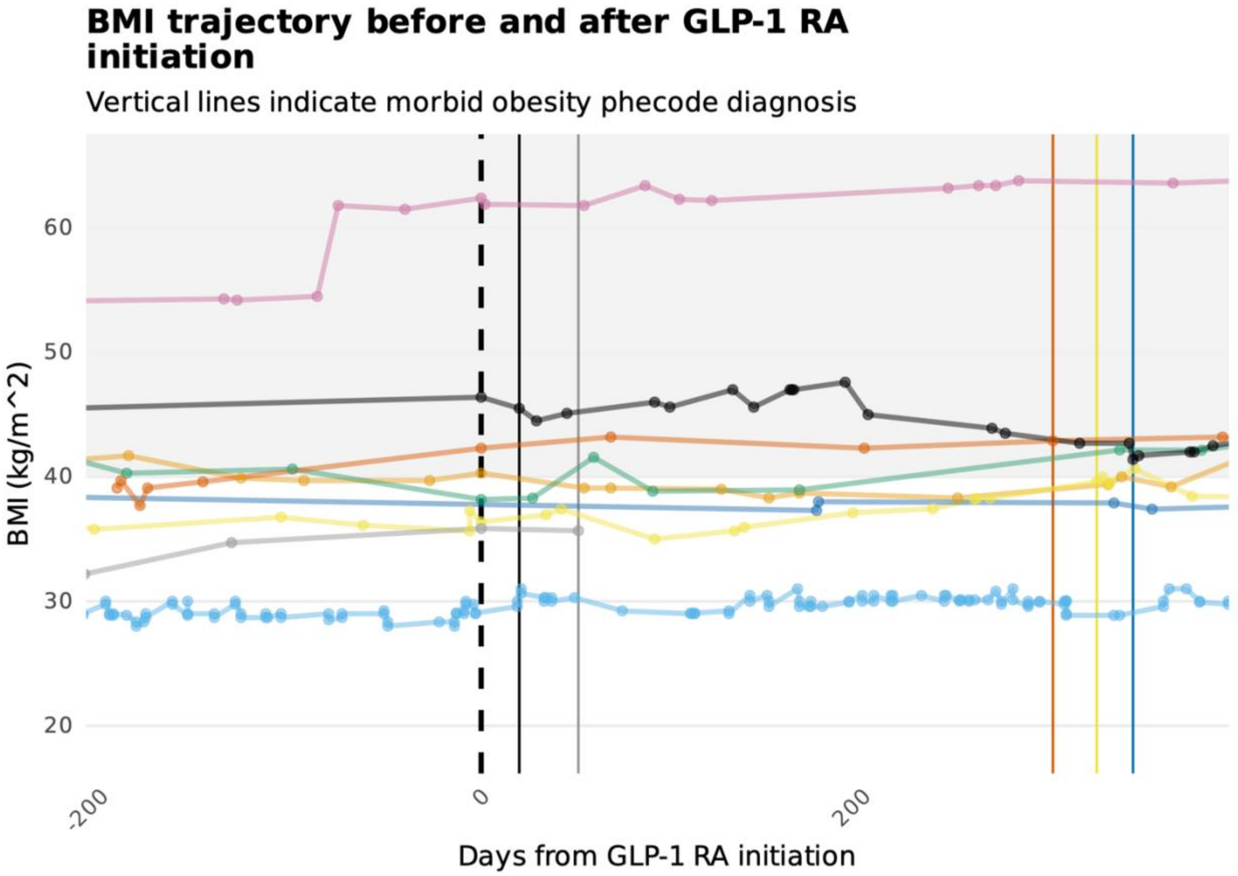


Figure S4. Random sample of 9 individuals who had a morbid obesity diagnosis after GLP-1 RA initiation and at least three BMI measurements within a year of treatment initiation. Some individuals were diagnosed with morbid obesity more than one year after treatment initiation.

Furthermore, we consulted clinicians specializing in metabolic and endocrine disorders who prescribe GLP-1 RAs. These clinicians confirmed that obesity-related ICD codes were rarely used, especially until the second generation incretin-based therapies became available. They also explained that clinicians may note an obesity-related code to enhance patients' access to GLP-1 RA.
